# Antimicrobial resistance patterns of bacterial isolates from bloodstream infections at Jinja regional referral hospital: A cross-sectional study

**DOI:** 10.1101/2023.08.09.23293917

**Authors:** Fahad Lwigale

**Affiliations:** Global Health Security Program, Infectious Diseases Institute, College of Health Sciences, Makerere University, Kampala, Uganda; Jinja Regional Referral Hospital, Jinja, Uganda

## Abstract

**Introduction:** Bloodstream infections are a commonly encountered clinical syndrome of public health concern with variable epidemiology. The characteristic of resistance to multiple antibiotics by their etiologic agents has limited the options for empirical sepsis case management. This study determined the most frequent aetiologic agents responsible for bloodstream infections, their antimicrobial susceptibility patterns, and infection sources in Eastern-Central Uganda.

**Materials and methods:** This retrospective study involved analysis of all non-duplicate blood culture reports from 2019 to 2021. The frequency and percentage of significant pathogenic organisms and contaminants were calculated. Positivity distribution including infection sources was determined. World Health Organization Network (WHONET) 2022 desktop software was used for antimicrobial susceptibility data analysis.

**Results:** The 1364 participants had a diagnostic yield of 114(6.8%) and a 1.5% contamination rate. Over 37% and 13% of infections were hospital and community-acquired respectively. Most etiologic agents were Gram-positive bacteria dominated by *Staphylococcus aureus* 39(34.2%). Gram-negatives mainly included unspecified *Coliforms* 12(10.5%), and *E. coli* 10(8.8%). Polymicrobial growth existed in 4(0.3%) cases. *S. aureus* was mainly resistant to Penicillin G (100%), Cefoxitin (60%), and Erythromycin (52.2%). Both *Escherichia coli* and unspecified *coliforms* showed high resistance to Ampicillin (87.5%) and third-generation Cephalosporins (100%). The least resistance was to Chloramphenicol and Carbapenems.

**Conclusion:** BSIs are mainly due to Gram-positive bacteria. Suspected sepsis cases by *S. aureus* in this region can be empirically managed using Gentamicin. Microbiology services should be routinely utilized to guide antimicrobial use, monitor blood culture contamination rates and resistance trends to strengthen regional antimicrobial stewardship.

## Introduction

Bloodstream Infections (BSIs) are some of the most commonly encountered and serious Healthcare-Associated Infections (HAIs) of public health concern(1-3). This results from microorganisms accessing the bloodstream where they reproduce and excrete toxins during septicemia (4). It is characterized by a number of symptoms including fever (>38°C), chills, hypotension, tachypnea, hypothermia (<36°C), apnea, or bradycardia depending on different factors including the age group (2, 5). A Bloodstream Infection (BSI) is confirmed by a positive blood culture in the microbiology laboratory whether primary or secondary (1, 2, 6). A primary BSI is a laboratory-confirmed positive blood culture without any link to another infected body site (5). A secondary BSI on the other hand involves the isolation of an organism within a 14-17-day period from the bloodstream with the same identity as one responsible for another existing body site-specific infection or contributes to a syndrome (5). This can be a urinary tract infection (UTI), respiratory tract infection (RTI), or skin and soft tissue infection (SSTI) among others.

Among all ages, BSIs are one of the major causes of morbidity and mortality in both developed and developing countries (7, 8). In the United States of America (USA), they are the 11th leading cause of mortality and among the top seven causes of death in Europe (9). Among febrile neutropenic patients, 6% of deaths are due to BSIs (10). They cause 13-15% of deaths in case of neonatal sepsis and 30-50% of deaths in developing countries (7). The mortality rate is 33.3% -46.9% in Italy (2). Among cancer patients, BSIs are responsible for a death rate of 24% and 33% in developed and then low- and middle-income countries (LMIC) respectively (11). As an emergency condition, sepsis requires immediate detection and identification of the causative agent for proper treatment in the current era of the Antimicrobial Resistance (AMR) silent pandemic(1, 12). For this matter, blood culture testing is an important tool for the diagnosis of BSIs and AMR surveillance at large(4, 6, 13).

Pathogen Recovery from the blood cultures has been observed to vary in Asia and Africa with comparable levels ranging from 6.11% to 27.1% (3, 10, 14-16). It was as high as 71% in Ethiopia (17). East African levels are 11.71% to 14.2% (18, 19). Uganda has previously been reported to have a diagnostic yield of 4.6% with 2.6% for Jinja (13). However, up to 14.1% positivity has been observed among cancer patients (11). The majority of studies unanimously report males to be the major victims of the BSIs ranging from 60.4% to 65% (2, 4, 9, 20). The majority belonged to 18-88 years of age (9, 20). BSIs of hospital origin have been reported to range between 10% to 81.5% (2, 4, 8). Community-acquired BSIs have also been reported to be as high as 91.1% to 91.6% elsewhere(14, 21).

Geographic and epidemiological factors have resulted in differences in the profile of BSIs observed(2, 8, 9, 11, 12). Gram-negative organisms have been reported to be dominant in some areas (11, 12, 15, 19, 22). ESKAPE composed of *Enterococcus faecium*, *Staphylococcus aureus*, *Escherichia coli*, *Klebsiella pneumoniae*, *Acinetobacter baumannii*, *Pseudomonas aeruginosa*, and *Enterobacter species* altogether contributes up to 70% of the observed BSIs (23). This is dominated by *E. coli*, *K. pneumoniae*, *S. epidermidis*, and *S. aureus* (23). Many others are similar to that (9, 12). Other studies indicate Gram-positive bacteria especially *Staphylococcus species* and *Enterococcus species* to be of a higher frequency(3, 4, 17, 20). AMR priority pathogens including *Staphylococcus aureus* (54%), *Escherichia coli* (19%), *Salmonella species* (11%), *Streptococcus pneumoniae*, and *Klebsiella species* (5%) were previously reported as the most dominant in Uganda (13).

Variably high resistance levels have been reported for multiple antibiotics including Gentamicin, Cephalosporins, and Carbapenems among *Enterobacterales* and *Acinetobacter baumannii* (9, 11, 18). There is an exhibition of different resistance phenotypes with levels up to 28.9% of Methicillin-Resistant *S. aureus* (MRSA), 23.9% of Extended-spectrum beta-lactamase (ESBL) producers while the Carbapenem-resistant (CarbR) are up to 26.3% (2). In Uganda, up to 57.1% resistance to Imipenem is reported (11). The majority of isolates have been observed to be more susceptible to Meropenem, Amikacin, Piperacillin/Tazobactam, and Ciprofloxacin (9, 12, 17, 24). These infections are commonly empirically managed using a combination of broad-spectrum intravenously administered antibiotics such as Ceftriaxone and Metronidazole. However, empirical therapy has been reported to facilitate the emergence of Antimicrobial resistance(7, 25). In this case, microorganisms can thrive in an environment composed of an antimicrobial agent previously capable of killing it or preventing its reproduction. Bacterial isolates of this nature have been associated with bloodstream infections (BSIs) where they exhibit Multi-Drug Resistant (MDR) phenotypes such as the Methicillin Resistant *Staphylococcus aureus* (MRSA), Extended Spectrum Beta Lactamase (ESBL) producers and Carbapenem-resistant (CarbR) organisms (2, 7, 18). This increases the possibility of therapy failure, longer hospital-stay periods with increased risk of acquisition of more infections, high healthcare costs, and a higher risk of death (9, 10, 25).

Despite the setup of the Medicines and Therapeutic Committee (MTC) and the Infection Prevention and Control (IPC) committees at Jinja regional referral hospital (Jinja RRH) by the ministry of health (MoH) of Uganda and implementing partners such as the Infectious Diseases Institute (IDI), there is a shortage of congregated up-to-date information to guide the choice of antibiotics for empirical therapy to ensure better control of antimicrobial use and consumption in the region served by the facility. This is in addition to a shortage in microbiology services as this site is the only supplier in the region and yet it is also affected by other factors including a shortage of supplies and staff resulting in intermittent availability of information for targeted therapy of bloodstream infections. The ever-changing microbiological profile of BSIs and the lack of published literature about this region makes it necessary to routinely generate, analyze and utilize the local data about infectious syndromes. This study aimed at determining the levels and distribution of the main causative agents for BSIs at Jinja RRH, their antibiotic profiles and major sources of the infections. This was well achieved. It will in turn help physicians, MTC, IPC committees, and policymakers to formulate guidelines for the management of BSIs, better infection prevention and control, and improve the antimicrobial stewardship and surveillance activities in Jinja RRH and the Eastern-Central region of Uganda.

## Materials and methods

### Operational definition

A Bloodstream Infection (BSI) refers to a positive blood culture test due to isolation of a significant pathogenic microorganism/s from the suspected patient’s blood sample. The organism can be bacterial or fungal in nature.

### Study design, setting, and population

This was a retrospective cross-sectional study conducted in 2022 using data generated over a two-and-a-half-year period from January 2019 through June 2021. This period was shortly after the stabilization of microbiology service delivery in the region. The Corona Virus Disease-2019 (COVID-19) pandemic was experienced in the same time zone and significantly impacted health service delivery. The data was collected at Jinja regional referral hospital. This is a 500-bed capacity hospital located along Bax Street within the central town of Jinja city in the Eastern-Central part of Uganda. As a referral hospital, this facility serves a population of approximately 4.5 million people from within Jinja and the surrounding areas of over 11 districts including Iganga, Mayuge, Kamuli, Buikwe, Lugazi, Kayunga, and Mukono districts.

The facility is equipped with a microbiology laboratory where examinations of various samples are conducted for routine patient care and management. Blood culture was carried out using the BD-BACTEC™ FX40 (BacT/ALERT) automated system. Organism identification was carried out using conventional methods up to the genus and species levels. Antimicrobial susceptibility testing was carried out using the Kirby-Bauer disk diffusion method following the Clinical and Laboratory Standards Institute (CLSI) guidelines; CLSI-2018-M100-S28(26), CLSI-2020-M100-S30(27) and CLSI-2021-M100-S31 (28). There were different antibiotic categories tested as commonly used against different organisms. The Penicillins included Penicillin G (10 units), Ampicillin(10µg), and Piperacillin (100µg). Cephalosporins included Ceftriaxone (30µg), Cefotaxime (30µg), Cefuroxime(30µg), Ceftazidime(30µg) and Cefoxitin (30µg). Combinations included Amoxicillin/Clavulanate (20/10)µg and Trimethoprim/Sulfamethoxazole(Co-trimoxazole) (1.25/23.75)µg. Erythromycin(15µg) was tested among the Macrolides. Carbapenems involved Imipenem (10µg), Meropenem (10µg) and Ertapenem (10µg). Fluoroquinolones mainly included Ciprofloxacin(5µg). The Lincosamide tested was Clindamycin (2µg). Aminoglycosides included Gentamicin (10µg) and Amikacin (30µg). Tetracycline and Chloramphenicol of composition 30µg each were also included.

This study aimed to determe the level of bloodstream infections and their common etiological agents; the percentage resistance and/or sensitivity of the etiological agents to common antibiotics; and the levels of community and healthcare-associated infections. A sample size of 410 participants was estimated using a 5% prevalence and a precision of 0.021 at 95% confidence. However, data from all patients (1364) with suspected BSI and non-duplicate blood culture tests carried out during the study period was considered. The higher sample size was aimed at increasing the chances of obtaining the minimum of thirty (30) isolates necessary to generate reliable cumulative antibiograms according to CLSI-M39A4E-2014 (29).This together was to increase precision and have a better representation of the population.

### Data Management

The data necessary for the study was accessed in 2022 and extracted from the African Laboratory Information System (ALIS) into an excel sheet. This was cleaned and anonymized by removal specific identifiers such as patient names and numbers before being analyzed. All sex types and age groups such as pediatrics, adolescents, and adults were included in the analysis. Only the first isolate of every patient in the study time period was included in the analysis to generate the antimicrobial susceptibility patterns as guided by CLSI-M39A4E-2014 (29). Organisms commonly found on the skin such as *Coagulase negative staphylococcus*(CoNS), *Corynebacterium Species*, and *Bacillus species* other than *B. anthracis* were treated as contaminants. The rest of the organisms were considered significant aetiologic agents when calculating the frequencies and respective percentages.

Hospital and community-acquired infections were defined based on the duration of patient hospitalization at the facility before sample collection. The infections diagnosed among personnel admitted and exposed to the hospital premises for at least 48 hours (two days) were treated as healthcare-associated (Hospital-acquired/Nosocomial) infections. Community-acquired infections on the other hand involved personnel exposed to the premises of the hospital for less than 48 hours. Respective frequencies and percentages were calculated.

### Ethics Statement

The Jinja Hospital Research and Ethics Committee approved the study with registration number **JREC 159/2022.** The data was obtained from records of personnel who seek medical services at the facility. This is kept in medical records including patients files, laboratory request forms and information systems for proper care and later reference. The Microbiology Laboratory records were anonymously analyzed for this study and no consent was obtained from individual personnel.

### Statistical analysis

Microsoft Excel (2016) was used for data cleaning and descriptive analysis. World Health Organization Network (WHO-NET 2022) desktop software was used to analyze the antimicrobial susceptibility data. Only the first isolate of every participant was involved in generation of antimicrobial susceptibility profiles. These were presented as percentage resistance and/or sensitivity/susceptibility. Results including positivity distributions, hospitalization history, organisms/aetiologic agents and AST data were presented in the form of frequencies and percentages in different tables and figures. There was a description of their distribution in socio-demographics including age, sex, and location (wards).

## Results

A total of 1364 participants were involved in the analysis. 800(58.7%) were male while females were 564(41.3%). The majority (86.2%) of participants were less than 12 years of age. A total of 114(8.4%) blood samples were positive. Of these, 93 had significant growth representing a true diagnostic yield of 6.8% as shown in **Fig 1**.

**Fig 1.**
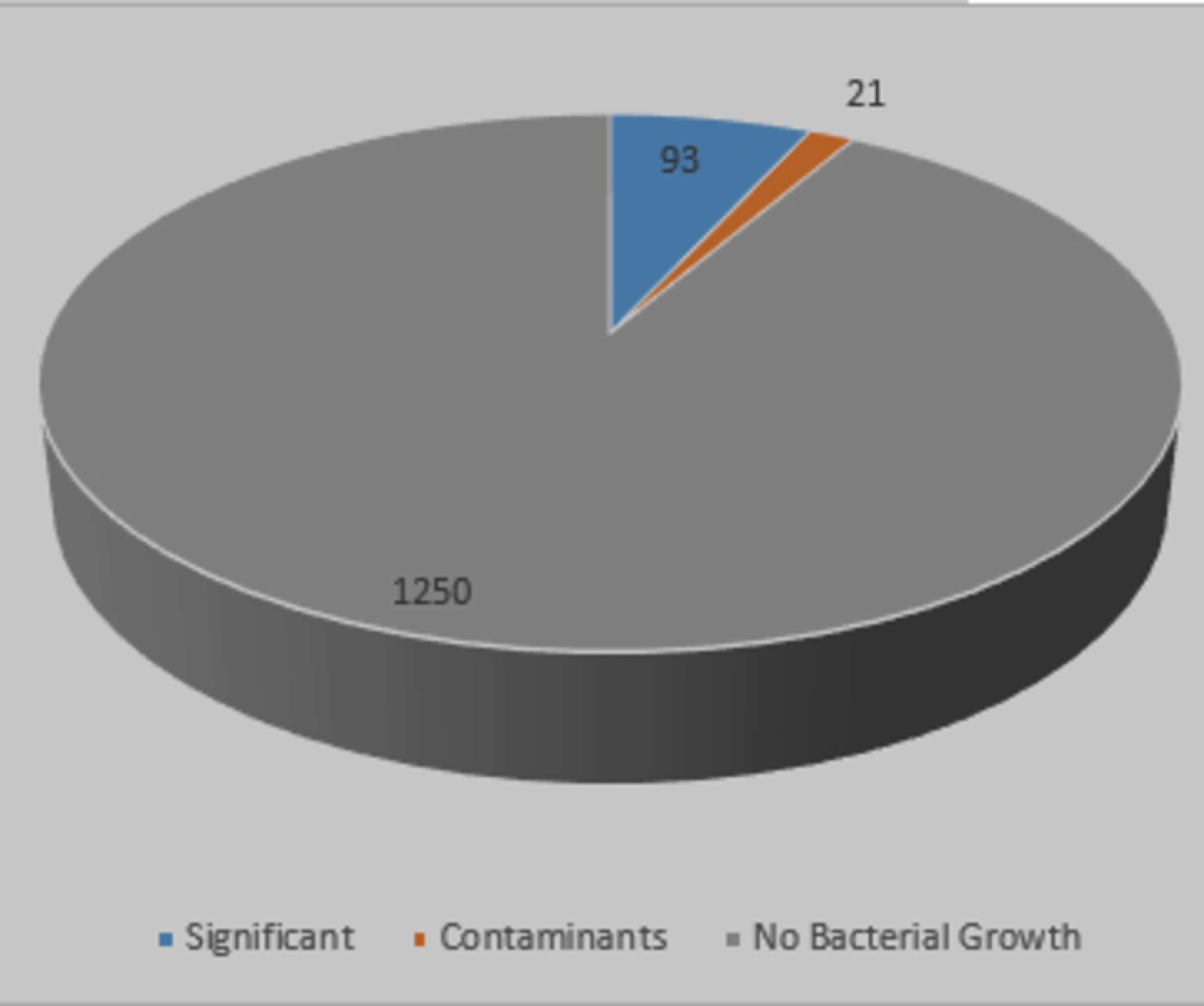
Levels of positivity for blood cultures. The observed prevalence of bloodstream infections is 6.8%.

The contamination rate observed was 1.5%. (Refer to **Table 1** for details). Positivity was observed more among the males 67(58.8%) than the females 47(41.2%) (**Table 1**). The frequency and percentage of isolated organisms are illustrated in **Fig 2**. Of the 114 positive cases, 68(60.0%) were Gram-positive bacteria while 44(38.6%) were Gram-negative. *Candida species* represented the remainder.

**Fig 2.**
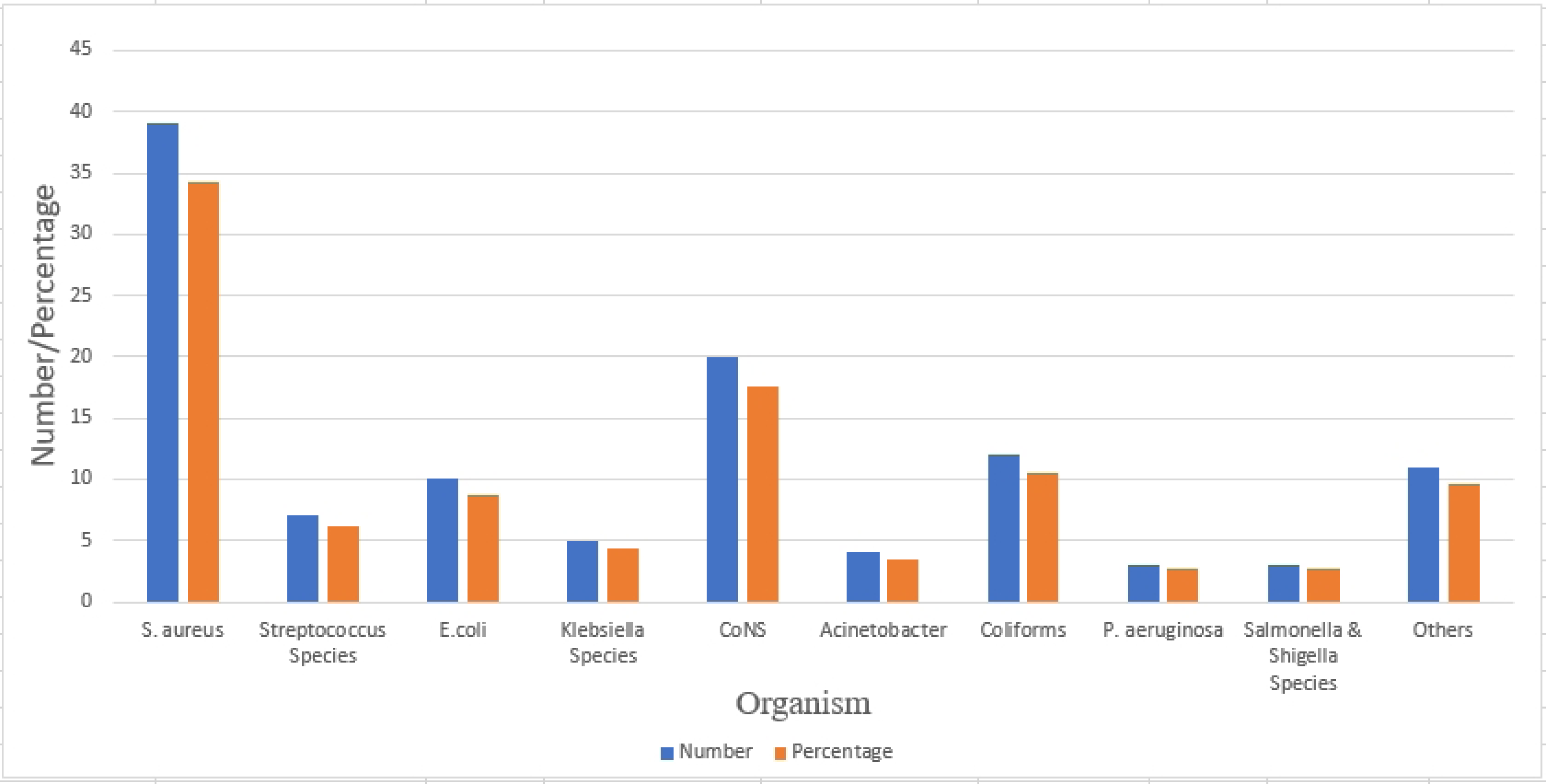
Frequency and Percentage of Isolated Organisms.

**Table 1.** Positivity distribution in the different population groups.

| Growth |  | Significant | Contaminant | NBG | Total |
| --- | --- | --- | --- | --- | --- |
| Frequency | n | 93 | 21 | 1250 | 1364 |
| Percentage | % | 6.8 | 1.5 | 91.6 | 100 |
| **Gram Reaction**<br>**n(%)** | Positive | 49(70) | 21(30) | N/A | 70 |
|  | Negative | 44(100) | 0(0) | N/A | 44 |
| **Sex n(%)** | Male | 54(6.8) | 13(1.6) | 733(91.6) | 800 |
|  | Female | 39(6.9) | 8(1.4) | 517(91.7) | 564 |
| **Age Group n(%)** | ≤12 | 78(6.6) | 18(1.5) | 1080(91.8) | 1176 |
|  | 13-18 | 4(9.1) | 0(0) | 40(90.9) | 44 |
|  | 19-35 | 2(3.3) | 0(0) | 58(96.7) | 60 |
|  | 36-59 | 5(9.4) | 2(3.8) | 46(86.8) | 53 |
|  | ≥ 60 | 4(12.9) | 1(3.2) | 26(83.9) | 31 |
| **Ward n(%)** | OPD | 1(4.3) | 0(0) | 22(95.7) | 23 |
|  | A&E | 3(7.7) | 0(0) | 36(92.3) | 39 |
|  | Pediatric | 75(6.6) | 18(1.6) | 1043(91.8) | 1136 |
|  | Medical | 3(5.9) | 2(3.9) | 46(90.2) | 51 |
|  | ICU | 0(0) | 0(0) | 8(100) | 8 |
|  | Others | 11(10.3) | 1(0.9) | 95(88.8) | 107 |
| **Total** |  | 93(6.8) | 21(1.5) | 1250(91.6) | 1364 |

A&E, Accidents and Emergency; ICU, Intensive Care Unit; NBG, No Bacterial Growth; N/A, Not Applicable; OPD, Outpatient Department

Gram positive bacteria majorly included, *Staphylococcus aureus* 39(34.2%), *Coagulase negative staphylococcus* 20(17.5%) then *Streptococcus species* 7(6.1%). Gram-negative bacteria on the other hand were made of general unspecified *Coliforms* 12(10.5%), *E. coli* 10(8.8%), *Klebsiella species* 5(4.4%), *Acinetobacter species* 4(3.5%), *Pseudomonas aeruginosa* 3(2.6%), then *Salmonella species* 2(1.8%). Other organisms included *Candida albicans*, *Enterobacter*, *Serratia*, *Citrobacter*, and *Shigella species*. Most cases were observed in the pediatric unit among children less than 13 years of age. Polymicrobial growth was observed in 4(0.3%) cases of which two involved a *Serratia species* and one had a *Bacillus species* in addition to the co-isolates.

Percentage resistance among *S. aureus* isolates is shown in **Fig 3**. Among *E. coli* and unspecified *coliform* isolates combined(n=22), Percentage resistance to Ampicillin, Cefuroxime, Ceftazidime, Cefotaxime, Gentamicin, Ciprofloxacin, Imipenem, Ertapenem, Co-trimoxazole, Tetracycline, and Chloramphenicol was 87.5%, 100%, 100%, 100%, 50%, 50%, 20%, 33.3%, 66.7%,60%, and 28.6% respectively. *Non-Enterobacterales* (*P. aeruginosa* and *A. baumannii*) together(n=7) had the following resistance profile; Ceftazidime (20%), Aztreonam (50%), Imipenem (0%), Meropenem (50%), Gentamicin (40%), Ciprofloxacin (33.3%) and Tetracycline (0%).

**Fig 3:**
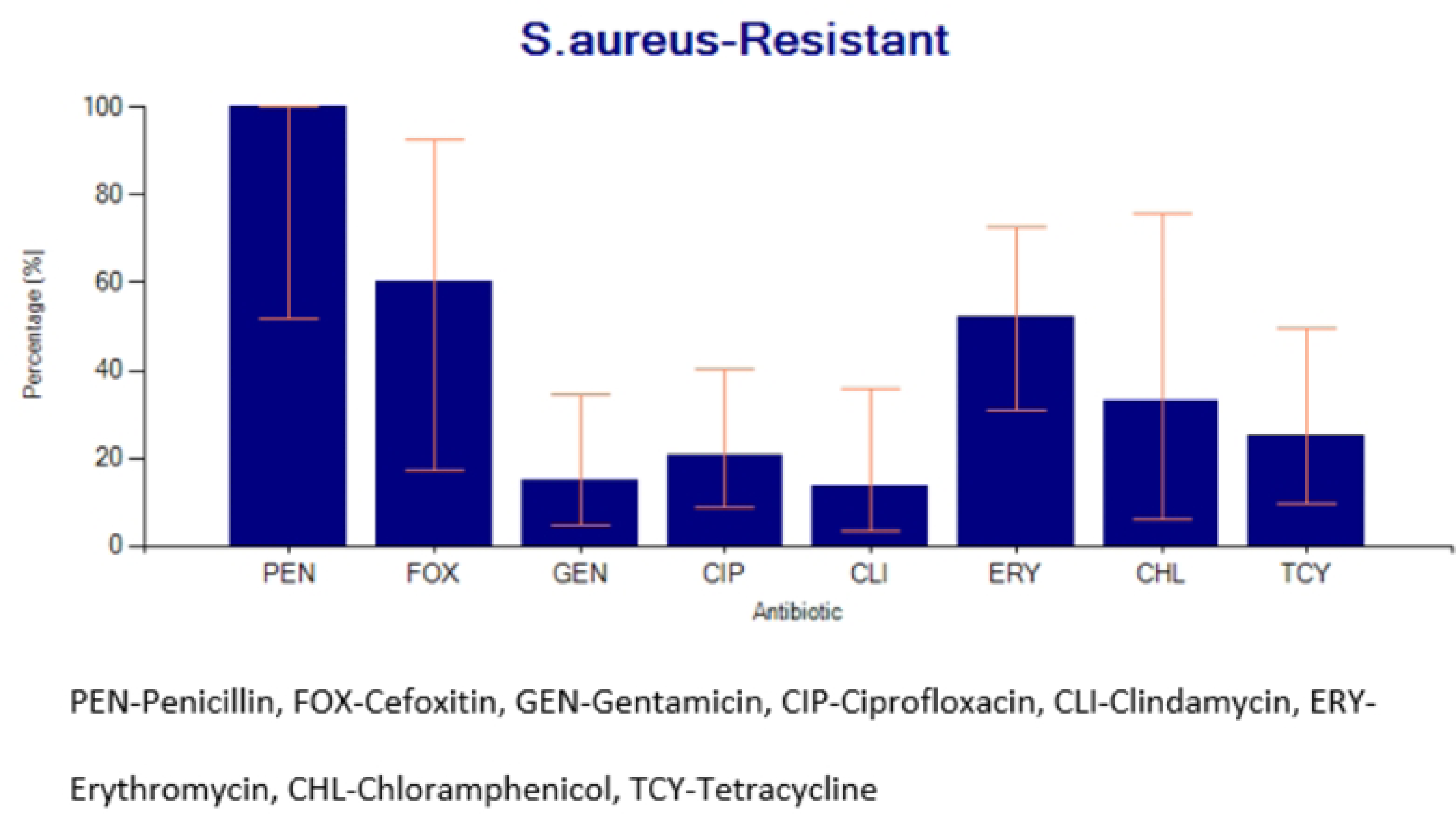
Percentage resistance for *Staphylococcus aureus* to common antibiotics.

**Table 2** shows the total number of different isolates and respective proportions tested against the antibiotics. Further, respective frequencies for the Resistant(R), Intermediate(I), and Susceptible(S) categories as outcomes of the antimicrobial susceptibility test (AST) reporting format have been revealed. This details the ratio of organism to antibiotic tested with respective outcomes compared with the number of organisms isolated.

**Table 2.** Frequency and percentage of organisms tested against different antibiotics.

| Antibiotic | AST<br>Result | Name of Organism |  |  |  |  |  |  |  |  |  |  |  |  |  |  |  |  |  |
| --- | --- | --- | --- | --- | --- | --- | --- | --- | --- | --- | --- | --- | --- | --- | --- | --- | --- | --- | --- |
|  |  | S. aureus<br>n =39 |  | Strepto<br>coccus<br>Species<br>n =7 |  | Coliforms<br>(Unspecified)<br>n =12 |  | E. coli<br>n =10 |  | Klebsiella<br>Species<br>n =5 |  | Acinetobacter<br>Species<br>n =4 |  | P. aeruginosa<br>n =3 |  | Salmonella<br>& Shigella<br>Species<br>n =3 |  | Others<br>n = 11 |  |
|  |  | n | % | n | % | n | % | n | % | n | % | n | % | n | % | n | %<sup>a</sup> | n | % |
| Ampicillin | R | - | - | - | - | 2 | 16 | 5 | 50 | 3 | 60 | - | - | - | - | 2 | 60 | - | - |
|  | I | - | - | - | - | 0 | 0 | 0 | 0 | 0 | 0 | - | - | - | - | 0 | 0 | - | - |
|  | S | - | - | - | - | 1 | 8 | 0 | 0 | 0 | 0 | - | - | - | - | 0 | 0 | - | - |
| Penicillin G | R | 6 | 15 | 0 | 0 | - | - | - | - | - | - | - | - | - | - | - | - | - | - |
|  | I | 0 | 0 | 0 | 0 | - | - | - | - | - | - | - | - | - | - | - | - | - | - |
|  | S | 0 | 0 | 1 | 14 | - | - | - | - | - | - | - | - | - | - | - | - | - | - |
| Piperacillin | R | - | - | - | - | - | - | 1 | 10 | 1 | 10 | - | - | - | - | - | - | - | - |
|  | I | - | - | - | - | - | - | 0 | 0 | 0 | 0 | - | - | - | - | - | - | - | - |
|  | S | - | - | - | - | - | - | 0 | 0 | 0 | 0 | - | - | - | - | - | - | - | - |
|  | R | 3 | 8 | - | - | - | - | - | - | - | - | - | - | - | - | - | - | - | - |
| Cefoxitin | I | 0 | 0 | - | - | - | - | - | - | - | - | - | - | - | - | - | - | - | - |
|  | S | 2 | 5 | - | - | - | - | - | - | - | - | - | - | - | - | - | - | - | - |
| Cefuroxime | R | - | - | - | - | - | - | 2 | 20 | - | - | - | - | - | - | - | - | 1 | 9 |
|  | I | - | - | - | - | - | - | 0 | 0 | - | - | - | - | - | - | - | - | 0 | 0 |
|  | S | - | - | - | - | - | - | 0 | 0 | - | - | - | - | - | - | - | - | 0 | 0 |
| Ceftazidime | R | - | - | - | - | - | - | 4 | 40 | 1 | 20 | - | - | 0 | 0 | - | - | 2 | 18 |
|  | I | - | - | - | - | - | - | 0 | 0 | 0 | 0 | 1 | 25 | 0 | 0 | - | - | 0 | 0 |
|  | S | - | - | - | - | - | - | 0 | 0 | 0 | 0 | 0 | 0 | 2 | 60 | - | - | 0 | 0 |
| Cefotaxime | R | - | - | - | - | 2 | 16 | 2 | 20 | 1 | 20 | - | - | - | - | 1 | 30 | - | - |
|  | I | - | - | - | - | 0 | 0 | 0 | 0 | 0 | 0 | - | - | - | - | 0 | 0 | - | - |
|  | S | - | - | - | - | 0 | 0 | 0 | 0 | 0 | 0 | - | - | - | - | 0 | 0 | - | - |
| Ceftriaxone | R | - | - | - | - | - | - | 2 | 20 | 2 | 40 | - | - | - | - | - | - | 1 | 9 |
|  | I | - | - | - | - | - | - | 0 | 0 | 0 | 0 | - | - | - | - | - | - | 0 | 0 |
|  | S | - | - | - | - | - | - | 0 | 0 | 0 | 0 | - | - | - | - | - | - | 0 | 0 |
| Gentamicin | R | 4 | 10 | 0 | 0 | 3 | 25 | 3 | 30 | 2 | 40 | 0 | 0 | 2 | 60 | - | - | 1 | 9 |
|  | I | 3 | 8 | 0 | 0 | 0 | 0 | 0 | 0 | 0 | 0 | 0 | 0 | 0 | 0 | 0 | 0 | 1 | 9 |
|  | S | 20 | 51 | 1 | 14 | 2 | 16 | 4 | 40 | 2 | 40 | 2 | 50 | 1 | 30 | - | - | 3 | 27 |
| Amikacin | R | - | - | - | - | - | - | 0 | 0 | - | - | - | - | - | - | - | - | - | - |
|  | I | - | - | - | - | - | - | 0 | 0 | - | - | - | - | - | - | - | - | - | - |
|  | S | - | - | - | - | - | - | 1 | 10 | - | - | - | - | - | - | - | - | - | - |
| Ciprofloxacin | R | 6 | 15 | 0 | 0 | 6 | 50 | 3 | 30 | 2 | 40 | 2 | 50 | 0 | 0 | 0 | 0 | 2 | 18 |
|  | I | 7 | 18 | 0 | 0 | 0 | 0 | 0 | 0 | 0 | 0 | 0 | 0 | 0 | 0 | 0 | 0 | 3 | 27 |
|  | S | 16 | 41 | 1 | 14 | 3 | 25 | 6 | 60 | 3 | 60 | 1 | 25 | 3 | 100 | 3 | 100 | 1 | 9 |
| Erythromycin | R | 12 | 31 | 4 | 57 | - | - | - | - | - | - | - | - | - | - | - | - | 1 | 9 |
|  | I | 2 | 5 | 1 | 14 | - | - | - | - | - | - | - | - | - | - | - | - | 0 | 0 |
|  | S | 9 | 23 | 2 | 57 | - | - | - | - | - | - | - | - | - | - | - | - | 0 | 0 |
| Meropenem | R | - | - | - | - | - | - | - | - | - | - | 0 | 0 | 2 | 60 | - | - | - | - |
|  | I | - | - | - | - | - | - | - | - | - | - | 0 | 0 | 0 | 0 | - | - | - | - |
|  | S | - | - | - | - | - | - | - | - | - | - | 1 | 25 | 1 | 30 | - | - | - | - |
| Imipenem | R | - | - | - | - | 2 | 16 | 0 | 0 | 0 | 0 | 0 | 0 | - | - | - | - | 0 | 0 |
|  | I | - | - | - | - | 1 | 8 | 0 | 0 | 0 | 0 | 0 | 0 | - | - | - | - | 0 | 0 |
|  | S | - | - | - | - | 2 | 16 | 4 | 40 | 4 | 80 | 1 | 25 | - | - | - | - | 2 | 18 |
| Etarpenem | R | - | - | - | - | 1 | 8 | 0 | 0 | 0 | 0 | - | - | - | - | 1 | 30 | 0 | 0 |
|  | I | - | - | - | - | 0 | 0 | 0 | 0 | 0 | 0 | - | - | - | - | 0 | 0 | 0 | 0 |
|  | S | - | - | - | - | 0 | 0 | 2 | 20 | 1 | 20 | - | - | - | - | 0 | 0 | 1 | 9 |
| Tetracycline | R | 5 | 13 | 0 | 0 | 2 | 16 | 1 | 10 | 1 | 20 | 0 | 0 | - | - | 1 | 30 | 1 | 9 |
|  | I | 4 | 10 | 1 | 14 | 1 | 8 | 0 | 0 | 0 | 0 | 0 | 0 | - | - | 0 | 0 | 0 | 0 |
|  | S | 11 | 28 | 1 | 14 | 1 | 8 | 0 | 0 | 0 | 0 | 2 | 50 | - | - | 0 | 0 | 0 | 0 |
| Clindamycin | R | 3 | 8 | 2 | 28 | - | - | - | - | - | - | - | - | - | - | - | - | - | - |
|  | I | 2 | 5 | 0 | 0 | - | - | - | - | - | - | - | - | - | - | - | - | - | - |
|  | S | 17 | 44 | 1 | 14 | - | - | - | - | - | - | - | - | - | - | - | - | - | - |
| Chloramphenicol | R | 2 | 5 | 0 | 0 | 1 | 8 | 1 | 10 | 3 | 60 | - | - | - | - | - | - | 3 | 27 |
|  | I | 0 | 0 | 1 | 14 | 0 | 0 | 0 | 0 | 0 | 0 | - | - | - | - | - | - | 0 | 0 |
|  | S | 4 | 10 | 1 | 14 | 1 | 8 | 4 | 40 | 0 | 0 | - | - | - | - | - | - | 1 | 9 |
| Co-trimoxazole | R | - | - | - | - | 0 | 0 | 2 | 20 | 2 | 40 | - | - | - | - | 0 | 0 | 2 | 18 |
|  | I | - | - | - | - | 0 | 0 | 0 | 0 | 0 | 0 | - | - | - | - | 0 | 0 | 0 | 0 |
|  | S | - | - | - | - | 1 | 8 | 0 | 0 | 0 | 0 | - | - | - | - | 1 | 30 | 1 | 9 |
| Vancomycin | R | - | - | 0 | 0 | - | - | - | - | - | - | - | - | - | - | - | - | 0 | 0 |
|  | I | - | - | 0 | 0 | - | - | - | - | - | - | - | - | - | - | - | - | 0 | 0 |
|  | S | - | - | 5 | 71 | - | - | - | - | - | - | - | - | - | - | - | - | 1 | 9 |
|  | R | - | - | - | - | - | - | - | - | 1 | 20 | - | - | 1 | 30 | - | - | 1 | 9 |
| Aztreonam | I | - | - | - | - | - | - | - | - | 0 | 0 | - | - | 0 | 0 | - | - | 0 | 0 |
|  | S | - | - | - | - | - | - | - | - | 0 | 0 | - | - | 0 | 0 | - | - | 0 | 0 |

AST, Antimicrobial Susceptibility Test; I, Intermediate; R, Resistant; S, Susceptible
<sup>a</sup>Percentages were calculated for isolates tested relative to the number of isolated organisms

**Table 3** shows the possible sources of infection in the different population groups for positive cultures. Altogether, 36.8% and 13.2% of the positive cases were hospital-acquired and community-acquired infections respectively. On a sad note, over 50% of the cases were of unknown origin.

**Table 3.**
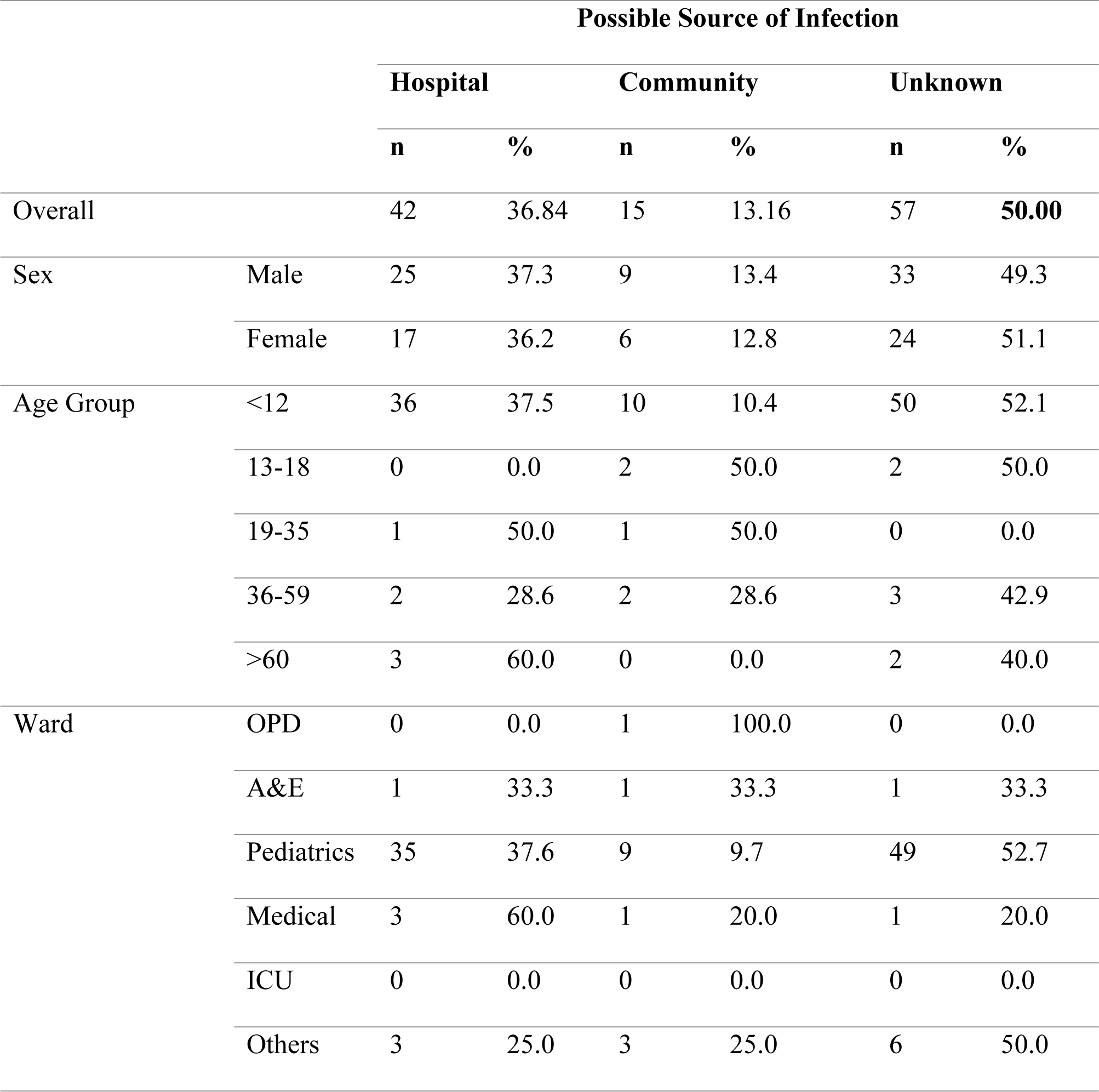
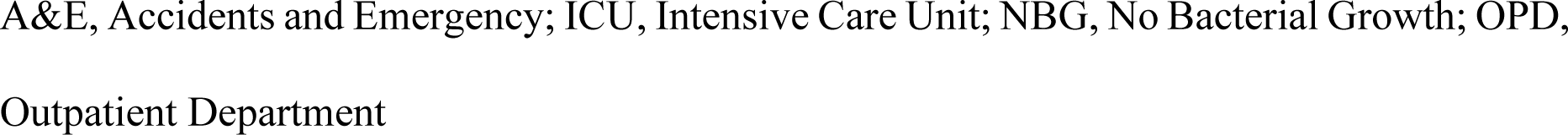
Likely infection source in the different population groups for positive cultures.

|  |  | Possible Source of Infection |  |  |  |  |  |
| --- | --- | --- | --- | --- | --- | --- | --- |
|  |  | Hospital |  | Community |  | Unknown |  |
|  |  | n | % | n | % | n | % |
| Overall |  | 42 | 36.84 | 15 | 13.16 | 57 | **50.00** |
| Sex | Male | 25 | 37.3 | 9 | 13.4 | 33 | 49.3 |
|  | Female | 17 | 36.2 | 6 | 12.8 | 24 | 51.1 |
| Age Group | <12 | 36 | 37.5 | 10 | 10.4 | 50 | 52.1 |
|  | 13-18 | 0 | 0.0 | 2 | 50.0 | 2 | 50.0 |
|  | 19-35 | 1 | 50.0 | 1 | 50.0 | 0 | 0.0 |
|  | 36-59 | 2 | 28.6 | 2 | 28.6 | 3 | 42.9 |
|  | >60 | 3 | 60.0 | 0 | 0.0 | 2 | 40.0 |
| Ward | OPD | 0 | 0.0 | 1 | 100.0 | 0 | 0.0 |
|  | A&E | 1 | 33.3 | 1 | 33.3 | 1 | 33.3 |
|  | Pediatrics | 35 | 37.6 | 9 | 9.7 | 49 | 52.7 |
|  | Medical | 3 | 60.0 | 1 | 20.0 | 1 | 20.0 |
|  | ICU | 0 | 0.0 | 0 | 0.0 | 0 | 0.0 |
|  | Others | 3 | 25.0 | 3 | 25.0 | 6 | 50.0 |

A&E, Accidents and Emergency; ICU, Intensive Care Unit; NBG, No Bacterial Growth; OPD, Outpatient Department

## Discussion

The BSIs are known life-threatening conditions with variable levels distributed. These are diagnosed when at least a single blood culture is carried out and a pathogenic microorganism is isolated. This study obtained a significant positivity level of 6.8%. This is greater than 4.6% and 2.6% for Uganda and Jinja respectively as earlier observed (13). This indicates an improvement in pathogen recovery from blood cultures. Similar levels have been obtained in other studies (14-16, 18). Variations of prevalence in different units have also been reported focusing on different facilities (3). Other previous studies obtained a relatively higher prevalence of up to 71% (17). This could be due to the study population of cancer patients majority of whom were neutropenic and hence at a higher risk for the BSIs (17, 30).

The highest prevalence of positivity was observed among the pediatrics. This could be explained by the low immune status of the children making them more vulnerable to the BSIs. Also, the largest number of samples were from children. This calls for the need to improve the infection prevention measures in these units. Routine blood culture is also recommended among adults fitting the definition to contribute more data about adults regarding BSIs. Up to 91.6% of the blood cultures had no bacterial growth. This could be due to the prior use of antibiotics between 1-28 days before blood culture testing as observed in many participants, especially in the period starting from early 2020.

More males (58.8%) were observed to have the BSIs and this is in agreement with the majority of previous studies(2, 4, 9, 20). Being male had also been associated with a high probability of having a BSI (crude OR 2.8, 95% CI 0.69–6.5; P=0.042) (17). Gram-positive bacteria (61.4%) were found to be more dominant compared to their Gram-negative counterparts (38.6%). Previous studies obtained similar results (3, 4, 13, 17, 20). Some research elsewhere also had differing observations where Gram-negatives were more than Gram-positives (11, 12, 15, 18, 19, 22). More variability was also shown in the study conducted in different ICUs (3). This provides a relatively larger tip of the iceberg regarding the variation of dominant organism distribution which calls for local data analysis and utilization.

*S. aureus* (34.2%) was the overall most commonly isolated organism at this site. The outcome was also observed elsewhere (4, 13, 17). Among Gram-negatives, it was majorly unspecified *Coliforms* (10.5%) followed by *E. coli* (8.8%) then *Klebsiella species* (4.4%), and *Pseudomonas aeruginosa* (2.6%). This is comparable to observations by other studies (12, 23, 31). *CoNS* were also more frequently isolated in the pediatric samples than *Streptococcus species* representing a contamination rate of 1.5% (21/1364). This was less than 13.6% and 4.8% observed at Princess Marina Hospital (PMH) and Tygerberg Hospital (TBH) neonatal units in Sub-Saharan Africa (21). Other studies have also reported higher rates (2, 10, 11, 16, 20, 23). This could be due to the high likelihood of contamination in children’s blood samples during sample collection. However, the observed contamination rate falls within acceptable limits ranging from 1-3% (6, 32). These isolates were not treated as true pathogens and no AST was done on them. This is a good practise in order to avoid unnecessary use of antibiotics in these children and their possible effects. On a sad note, only a single blood culture set was commonly done for the different individuals. This is most likely due to the limited availability of testing materials such as blood culture bottles among other factors. This limits the capability to confirm the fact that common commensals could be responsible for some of the true infections observed especially among immunocompromised individuals including children. This in turn leaves them vulnerable to the BSIs of that origin. However, it is recommended to employ appropriate policies to monitor and reduce the contamination levels to less than 1% and enable avoidance of the associated consequences(32).

Antimicrobial Susceptibility Testing (AST) was variably done based on the availability of testing materials. Each organism type was generally tested against at least 8 antibiotics except for *Pseudomonas aeruginosa* and *Acinetobacter species*. However, the ratio of the individual isolate to antibiotic tested was quite low (Refer to **Table 2**). This is similar to the observation at Mulago hospital (33). This clearly shows the need for Uganda to improve in terms of supply management and ensure the availability of necessary logistics for laboratory testing. For the number of *S. aureus* isolates tested against Cefoxitin, 60% turned out resistant indicating a possibility of high levels of MRSA in circulation. These (100%) being β-Lactamase producers, they had already shown no response to Penicillin G. However, Gentamicin 74.1% (%S 95% CI=53.4-88.1), Ciprofloxacin 55.2% (%S 95% CI=36.0-73.0), and Tetracycline 55% (%S 95% CI=32.0-76.2) showed better activity against *S. aureus* compared to other agents.

*E. coli* and unspecified *coliforms* combined (n=22) showed relatively high levels of resistance to commonly used antibiotics for empiric and targeted case management in the study setting such as the third-generation Cephalosporins. They also showed 20% and 33.3% resistance to Imipenem and Ertapenem respectively among the tested isolates. This indicates a possibility of increasing resistance to the reserved options limiting the availability of better alternatives when faced with cases of MDR pathogens. A similar feature was observed among the *Non-Enterobacterales* although they were susceptible to Tetracycline. However, there were fewer than 30 isolates for these two groups of organisms making it impossible to make appropriate conclusions about their susceptibility patterns as guided by Clinical and Laboratory Standards Institute (CLSI-M39A4E-2014) (29).

Data were scarce regarding the hospitalization of patients prior to sample collection. This was observed in over 50% of the data. However, based on the available information, at least 37% had their BSI being nosocomial. This is linked to the fact that they were exposed to the hospital environment for more than 48 hours before being sampled for testing. This increases the likelihood that the infection was acquired from the hospital making it necessary to strengthen IPC measures observed in the facility before it is too late. Similar observations were made elsewhere (2). However, higher levels of the same kind have been reported among non-ICU hospitalized patients (8). The remaining 13% of the positive cases can be linked to community-acquired infections. It is less than what is reported by other studies but there is room for more (14, 21). This calls for the need to improve information management at the testing and surveillance sites to enable capturing of all necessary data about the cases managed. Further study might also be needed to identify the possible causes of the data incompleteness at the different surveillance sites.

## Conclusions

*S. aureus* is the most common organism responsible for bloodstream infections at this site. There is a relatively high level of resistance to commonly used antibiotics among all organisms. Currently, Gentamicin can be used to empirically manage suspected *S. aureus* BSIs in this region. It is recommended to routinely utilize microbiology services with proper data management to guide antimicrobial use and monitor blood culture contamination rates and antimicrobial resistance trends to strengthen antimicrobial stewardship and surveillance policies in the Eastern-Central region of Uganda.

## Data Availability

The dataset to support the conclusions is published and freely accessible via the Dryad website (https://doi.org/10.5061/dryad.3bk3j9kqg)

## Acknowledgments

The author appreciates all contribution from the administration and staff of Jinja Regional Referral Hospital, especially the Children’s ward, Nalufenya for the great work together with Ms. Kasuswa Sophia, Mr. Kasibante Samuel, and Ms. Matinyi Sandra for the research encouragement. The author thanks the Infectious Diseases Institute for the support to offer microbiology services at this site.

